# Loss of function mutation in *ELF4* causes autoinflammatory and immunodeficiency disease in human

**DOI:** 10.1101/2021.09.22.21263689

**Authors:** Gan Sun, Luyao Qiu, Yunfei An, Yuan Ding, Lina Zhou, Junfeng Wu, Xuemei Tang, Huawei Xia, Lili Cao, Fuping You, Xiaodong Zhao, Hongqiang Du

## Abstract

Monogenic autoinflammatory diseases (mAIDs) are a heterogeneous group of diseases affecting primarily innate immunity, with various specific genetic causes. Genetic diagnosis of mAIDs can assist in the patient’s management and therapy. However, a large number of sporadic and familial cases remain genetically uncharacterized. Here, we described a pediatric patient suffering from recurrent viral and bacterial respiratory infection, refractory oral ulcer and constipation, who was clinically diagnosed of inborn errors of immunity (IEI). In an effort to establish genetic diagnosis, no known causative genes were identified by whole-exome sequencing. However, manually going through bioinformatically predicted candidate genes, we suspected and prioritized *ELF4* (chrX:129205133 A>G, c.691T>C, p.W231R) as the genetic cause for our patient. We then evaluated the pathogenicity of this mutation by both various bioinformatic methods and preliminary but definitive experimental approach. Our data suggested that W231R mutant ELF4 is a “loss of function” mutation causing decreased protein stability and decreased trans-activation activity. Thus, we identified a novel mAID, which we termed “X-linked autoinflammatory and immunodeficiency disease associated with ELF4, X-AIDE”.

## Introduction

Monogenic autoinflammatory diseases (mAIDs) are a heterogeneous subgroup of inborn errors of immunity (IEI) diseases or primary immunodeficiency diseases (PIDs), which are marked by hyperactivation of the innate immune system without apparent involvement of either autoantibodies or autoreactive T cells^1^. The autoinflammatory disease spectrum has expanded rapidly to include more than 40 distinct monogenic conditions, which can be roughly categorized into “Inflammasomopathies and other diseases arising through IL-1-family cytokines”, “Type I Interferonopathies”, “Disorders of NF-kB and/or aberrant TNF activity”, and “Autoinflammation mediated by other mechanisms”^2^. Across these categories, mAIDs vary widely in their presentation and clinical manifestations. While the definitive diagnosis is supported by genetic testing, in many practical settings, the diagnosis remains clinical^3^. Even in the monogenic diseases a clinical approach is first necessary to narrow the genetic search for the correct disease^4^. mAIDs should be suspected in patients with recurrent fever unexplained by infections and/or with episodic stereotypic symptoms in various organs and tissues, especially the skin, gastrointestinal tract, musculoskeletal system, eyes, mucous membranes and central nervous system. When a clinical diagnosis of mAID is made, a genetic approach should be taken^5^, preferably such as the next-generation whole exome sequencing (WES) if applicable, which has fueled the discovery of novel genes in the past several years.

By definition, mAIDs do not involve adaptive immune responses, however, this simplistic division is at best a first approximation. Innate and adaptive immunity are closely interconnected, and dysfunction in one often disturbs function in the other^2^. For example, some diseases now considered autoinflammatory feature autoantibodies. In some of the newly described diseases, recurrent infections, indicative of an immunodeficiency, are seen along with autoinflammatory-related symptoms^6^. Thus, a more accurate understanding of the pathogenesis of mAIDs may need a thorough investigation of both innate and adaptive immunity.

ELF4 belongs to the E-Twenty-Six (ETS) domain transcription factor family and is involved in a variety of biological processes, including adaptive immune response^7^. Previously, we have shown that ELF4 is a critical transcription factor for the host antiviral response, during which ELF4 cooperates with nuclear factor-kB (NF-κB) to induce robust interferons and inflammatory cytokine production^8^. In addition, we recently showed Elf4-deficient mice are highly susceptible to DSS-induced colitis, a subgroup of inflammatory bowel disease^9^. The potential role of ELF4 in the pathogenesis of “Type I Interferonopathies” or “Disorders of NF-κB and/or aberrant TNF activity” has not been revealed in human.

Here we studied a Chinese family with a X chromosome-linked disorder demonstrating as recurrent viral and bacterial respiratory infection, refractory oral ulcer and constipation. Whole-exome sequencing on the proband and his parents was negative for any known causative genes. However, manually going through bioinformatically predicted candidate genes, we suspected and prioritized *ELF4* (chrX:129205133 A>G, c.691T>C, p.W231R), which was Sanger sequencing confirmed first, as the genetic cause for our patient. We then evaluated the pathogenicity of this mutation by both various bioinformatic methods and preliminary but robust experimental approach. Our data suggested that W231R mutant ELF4 is a “loss of function” mutation causing decreased protein stability and decreased trans-activation activity. Thus, we identified a novel mAID, for which we proposed the term “X-linked autoinflammatory and immunodeficiency disease associated with ELF4, X-AIDE”.

## Results

### Case characteristic summary

The patient (P1) presented with frequent upper and lower respiratory tract (URT) infection with blood routine test mostly showing normal WBC but increased lymphocyte percentage, which indicated a viral infection. He also suffers from recurrent and refractory oral ulcer and refractory constipation but no hematochezia, melena, abdominal pain, fever of unknown nor diarrhea. Regular IVIG infusion at a supportive dose ranging from 400mg/kg to 1g/kg was advised and strictly followed by the patient. Frequency of respiratory infection dropped dramatically, however, oral ulcer still bothers him at a slightly lower frequency. The patient is at school age now. (Detailed medical history is available from the corresponding authors upon reasonable request).

### Genetic findings

Upon suspecting PID, we performed whole-exome sequencing of whole blood sample from the proband and his parents. A hemizygous *ELF4* mutation (chrX:129205133 A>G, c.691T>C, p.W231R) drew our attention. Sanger sequencing confirmed this (Fig. 1A), which is consistent with their phenotype. Firstly, this mutation does not occur in the 1000g database, ExAC database and in-house database. This amino acid site is extremely conserved among the species (GERP++ score 5.45, classified as “conserved”) and among ETS domain transcription factor super family members (Fig. 1B). Pathogenicity prediction using multiple methods classified this mutation as “Damaging”, “Probably damaging”, “Disease causing”, “Pathogenic” or “Functional Impact High” (Table 2)^11^.

**Figure 1.**
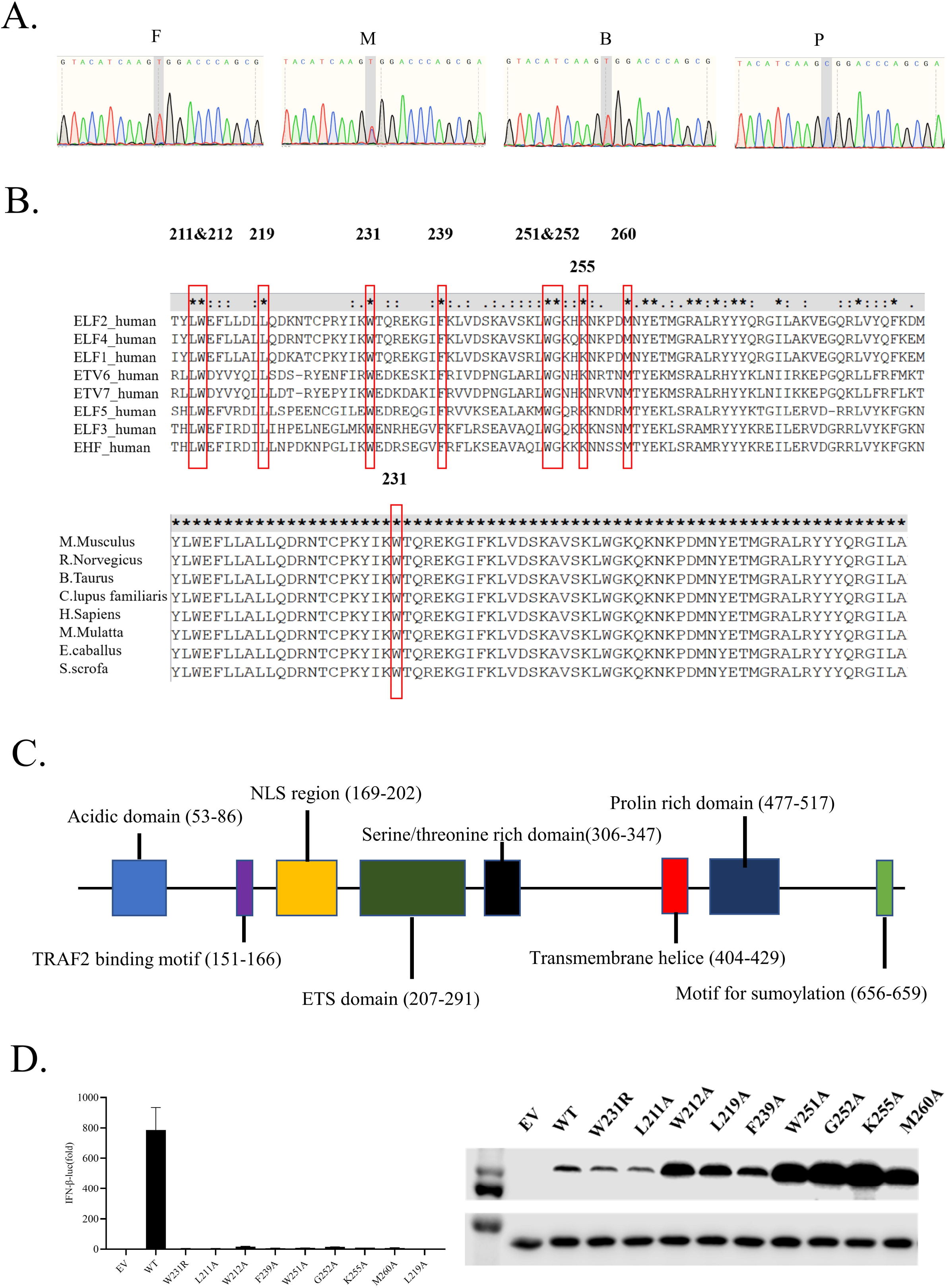
A missense mutation in *ELF4* is identified in a patient with recurrent respiratory infection and oral ulcer. A. Sanger sequencing of the ELF4 gene in P1 and family members. Genomic DNA was used. B. Amino acids sequence from ETS domain transcription factors family (upper) and ELF4 of different species are aligned. Some highly conserved sites are indicated. C. Schematic diagram of the human ELF4 protein. D. IFN-β dual luciferase reporter screening for essential sites of ELF4 in ETS domain for induction of type I interferon (left) and western blot confirming the expression of plasmids (right).

Secondly, ELF4 is a transcription factor and plays critical roles in both innate and adaptive immune responses^8^. This mutation site locates at the ETS domain of ELF4 (Fig. 1C), which is critical for its DNA binding^13^.

Thirdly, when we test the significance of highly conserved amino acid sites in the ETS domain of ELF4 in the induction of type I interferons (Fig. 1D), we found site 231 is among those essential for type I interferon induction (Fig. 1D). Taken together, we speculated that ELF4 mutation is the genetic cause for this undefined PID.

### W231R is a “loss of function” mutation

To explore the more specific effects of this mutation, we first examined the stability change of the mutant ELF4 using DynaMut, a widely cited web-based tool for analysis and prediction of protein stability changes upon mutation using Normal Mode Analysis^13^. Because there is no experimental crystal structure data for both human and mouse ELF4 protein, we here used AlphaFold2-predicted structure data in PDB format^14^. The predicted ΔΔG is -1.380 kcal/mol, classified as “Destabilizing”. Structurally, various atomic contacts and bonds are predicted to be disrupted by the mutation (Fig. 2A). The Δ vibrational entropy energy between wild-type and mutant indicated increase of molecule flexibility (ΔΔS_Vib_ ENCoM: 1.215 kcal.mol^-1^.K^-1^) (Fig. 2B). Consistently, we found decreased expression of mutant ELF4 in patient’s PBMC (Fig. 2C). To further confirm this, we constructed wild type and mutant ELF4 expression plasmids and performed Cycloheximide Chase Assay in HEK293T cells. Mutant ELF4 showed decreased stability compared to WT ELF4 (Fig. 2D).

**Figure 2.**
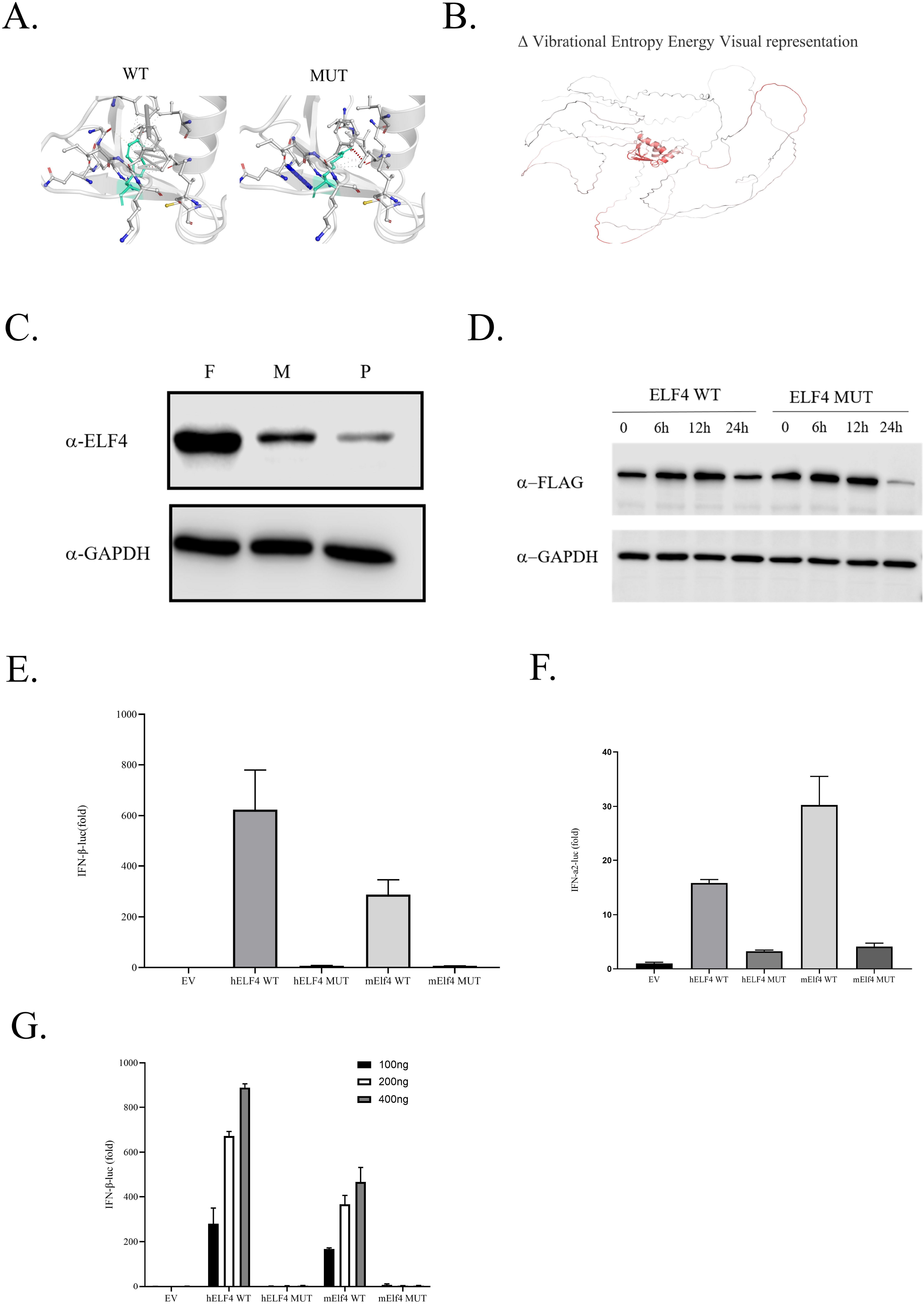
W231R mutation confers ELF4 decreased stability and trans-activation activity. A. Interactomic interactions changes predicted by DynaMut using AlphaFold2 predicted ELF4 structure. Wild-type and mutant residues are colored in **light-green** and are also represented as sticks alongside with the surrounding residues which are involved on any type of interactions. B. Visual representation of Δ Vibrational Entropy Energy predicted by DynaMut using AlphaFold2 predicted ELF4 structure. Amino acids colored according to the vibrational entropy change upon mutation. **BLUE** represents a rigidification of the structure and **RED** a gain in flexibility. C. Expression of ELF4 in PBMCs from patient (P), patient’s father (F) and patient’s mother (M). D. Protein stability test of mutant ELF4. Cycloheximide Chase Assay for W231R mutant ELF4 and WT control. Representative images from 2-3 independent experiments. E. IFN-β dual luciferase reporter assay for WT and mutant ELF4. hELF4, human ELF4; mELF4, mouse ELF4. Data are derived from 2-3 independent experiments. F. IFN-α dual luciferase reporter assay for WT and mutant ELF4. hELF4, human ELF4; mELF4, mouse ELF4. Data are derived from 2-3 independent experiments. G. IFN-β dual luciferase reporter assay for the dose-effect of WT and mutant ELF4. hELF4, human ELF4; mELF4, mouse ELF4. Data are derived from 2-3 independent experiments.

Next, we explored the intrinsic functional impact of this mutation first using MutationAssessor, a widely used tool for prediction of functional impact of amino-acid substitutions in proteins^15^. The Functional Impact score is 3.55 indicating “highly functionally affected” (Table 2). We further tested this in our type I interferon luciferase reporter system and showed that WT ELF4 (human and mouse) can activate IFN-β and IFN-α reporter in a dose-dependent manner while mutant ELF4 showed almost abolished trans-activation activity (Fig. 2E-2G). Collectively, our data suggested that W231R mutant ELF4 is a “loss of function” mutation in two senses, decreased protein stability and decreased trans-activation activity (possibly for decreased DNA binding activity).

## Discussion

Autoinflammatory diseases were first recognized over two decades ago as distinct clinical and immunological entities caused by dysregulation in the innate immune system^16^. Since then, advances in genomic techniques have made reaching a genetic diagnosis easier and led to the identification of new monogenic disorders. We here identified a novel mAID, which we termed “X-linked autoinflammatory and immunodeficiency disease associated with ELF4, X-AIDE”.

As pediatric immunologist, our diagnosis of a disease for a patient is firstly based on clinical manifestations and medical history of the patients and begins with a clinical suspicion or clinical diagnosis. For the diagnosis of mAIDs, a clinical suspicion should be made when: 1) Evidence for systemic inflammation (elevated acute phase reactants during disease flare e.g. C-reactive protein (CRP), erythrocyte sedimentation rate (ESR) and serum amyloid A (SAA). In some diseases, patients do not have strong evidence for systemic inflammation but may have organ or tissue specific inflammation (e. g. autoinflammatory disorders with prominent cutaneous manifestations where inflammation can be confirmed by skin biopsy). 2) The disease is likely to be inherited as a monogenic trait (early-onset symptoms in sporadic patients or multiple affected siblings or dominantly inherited symptoms). 3) Plausible candidate causal gene^3^. Our patient suffered from refractory oral ulcer with early onset and prolonged course. This local vasculitis is similar to what commonly seen in patients with Behçet’s disease or with Haploinsufficiency of A20 (HA20)^17^, which indicates this disease may belong to the category of “Disorders of NF-κB and/or aberrant TNF activity”. In addition, our patient also suffers from recurrent viral and bacterial respiratory infection with early onset and prolonged course, which further encourage us to made the diagnosis of primary immunodeficiency.

In some recently described novel mAIDs, autoinflammatory disease causing mutations also lead to defective immune responses and predispose to infection. Mutations in *PLCG2* cause autoinflammation with antibody deficiency^18^. Deficiency of components of the linear ubiquitin chain assembly complex (LUBAC) leads to severe autoinflammation with immunodeficiency in association with polyglucosan myopathy^19^. Mutations in *TRNT1* produce a syndrome of siderblastic anemia with B cell immunodeficiency, periodic fevers and developmental delay^20^. Patients with deficiency in WDR1 manifest autoinflammatory periodic fever, immunodeficiency and thrombocytopenia^21^. One potential explanation is that cell-type specific effects of mutated proteins lead directly to the combination of reduced immunity and enhanced inflammation. We previously showed ELF4 is critical for type I interferon induction during viral and bacterial infection^8^. Our patient harbors W231R mutation in *ELF4* gene (Fig. 1A and 1B), which located within the the ETS domain (Fig. 1C). This amino acid site is highly conserved among the species (Table 2) and among ETS domain transcription factor super family members (Fig. 1C). Mutant ELF4 showed almost abolished activity for type I interferon trans-activation (Fig. 2E-2G), which is consistent with the susceptibility of our patient to viral and bacterial infection. Decreased CD8 T cells and NK cells (Table 1), which is consistent with findings in Elf4 knockout mice^22^. The hyperinflammation demonstrated as refractory oral ulcer in our patient suggests that ELF4 may negatively regulate a proinflammation pathway, in which loss of function of ELF4 results in uncontrolled inflammation. In the preparation of our manuscript, a report with similar findings was published^23^. Two of their three patients harbor a W251S missense mutation, the other is a frameshift mutation, all of which cause loss of function of ELF4. In our screening assay, we also found amino acid 251 is essential for type I interferon induction (Fig. 1D). These three patients presented with fever, oral ulcers and mucosal inflammation, but not infection. In deed they found ELF4 promotes IL1RN (an anti-inflammation gene) and limits Trem1 expression in macrophages. Our patient lacks intestinal mucosal inflammation, but presents with constipation. The phenotype discrepancy between patients with W251S and our patient needs further investigation.

**Table 1.** Comprehensive analysis of peripheral lymphocytes subsets

**Table 2.** The prediction of the identified variant in *ELF4*

In conclusion, in the present study we described a patient presenting with recurrent viral and bacterial respiratory infection, refractory oral ulcer and constipation. After a clinical diagnosis of mAID is made, we further pursued the genetic diagnosis. Whole exome sequencing found no known genes for the patient but *ELF4* (chrX:129205133 A>G, c.691T>C, p.W231R) is a strong candidate novel gene. We then evaluated the pathogenicity of this mutation by both various bioinformatic methods and confirmed it with experimental approach. Thus, our patient presents as a novel mAID, which we termed “X-linked autoinflammatory and immunodeficiency disease associated with ELF4, X-AIDE”. More mechanistic studies are needed for more accurate understanding of this new disease.

## Data Availability

The data that support the findings of this study are available from the corresponding authors upon reasonable request.

## Materials and methods

### Patient

The ethics committee of Children’s Hospital of Chongqing Medical University approved the study. Blood samples from patients and unaffected relatives were collected for molecular studies, which were performed in accordance with the Declaration of Helsinki.

### Patient consent statement

Written informed consent for participation in the study and for the publication of the research work was obtained from patients’ parents.

### Genetic Analyses

Genomic DNA were isolated from peripheral blood samples. Disease-causing mutations were screened using whole genome sequencing (WES) (MyGenostics, Inc.). Candidate mutations were confirmed by Sanger sequencing using the following primers: seq F: 5’-AAGACCAAGGGCAACCGAAG -3’, seq R: 5’-CCATGAAAGCGAGGAGGACT -3’.

### Construction of plasmids

7.1-pCMV-3×Flag wild type human and mouse ELF4 plasmids were as previously described^8^. W231R mutant of human ELF4 and W230R mutant of mouse ELF4 were generated by overlap PCR method using WT as template. Mutations were confirmed by Sanger sequencing.

### Cells

HEK 293T (Human Embryonic Kidney 293 cells transformed by expression of the large T antigen from SV40) (ATCC, CRL-11268) were cultured in Dulbecco’s modified Eagle medium (DMEM).

### Western blotting

Cells were harvested and lysed in RIPA buffer (Beyotime, P0013C), containing complete mini protease inhibitor cocktail (Roche, 04693124001). Proteins were separated by SDS-PAGE and transferred to polyvinylidene fluoride membranes (GE, PVDF 0.45UM, 10600023). Non-specific binding was blocked using 5% non-fat milk and the primary antibody (anti-Flag, Sigma F3165, anti-GAPDH, Invitrogen MA1-16757, anti-ELF4, Sigma AV38028 was incubated at 4°C overnight, the secondary antibody was incubated at room temperature for 1 h. Signals were visualized by chemiluminescence (Millipore Corporation, Billerica, MA, USA).

### Cycloheximide Chase Assay

This assay is adapted from previous report^24^. Briefly, HEK293T cells were plated and transfected with various WT and mutant 7.1-pCMV-3×Flag expression plasmids. 24 hours later, cycloheximide was added (1ug/ml) and cells were collected at different time point for western blot detection.

### Statistical analysis

Samples were compared using two-tailed, unpaired Student’s *t*-test with GraphPad Prism 7.00. Error bars were represented by SEM. \**P* < 0.05, \*\**P* < 0.01, \*\*\**P* < 0.001.

## Authors’ contributions

HQ.D and XD.Z conceived and supervised the project, reviewed and revised the manuscript; HQ.D designed the experiments, analyzed data and wrote the manuscript. G.S performed the experiments, analyzed data and followed the patient. HW.X and LL.C provided experimental expertise. FP.Y provided scientific sights in designing the project and revising the manuscript. Others provided essential help for clinical management and follow-up of the patient and valuable clinical expertise in reviewing the manuscript.

## Acknowledgement

We thank the patient and his family for their cooperation, especially for the patient’s mother whose perseverance in caring for her child gives us strength to pursue a definitive genetic diagnosis and next a more effective personalized treatment.

## Fundings

Sanming Project of Medicine in Shenzhen (SZSM201812002);

Livelihood Project of Chongqing, China (cstc2018jscxmsybX0005)

National Natural Science Foundation of China (81974255)

## Author consent

We confirm that the manuscript has been read and approved by all named authors.

## Ethics approval

The ethics committee of Children’s Hospital of Chongqing Medical University approved the study.

Blood samples collection were performed in accordance with the Declaration of Helsinki.

