## Supplementary material for "Loss of function mutation in *ELF4* causes autoinflammatory and immunodeficiency disease in human": Tables

Table 1. Comprehensive analysis of peripheral lymphocytes subsets

| lymphocytes tested | | Percentage (%) | Percentage reference range | Absolute numbers (cells/ul) | Absolute numbers reference range |
| --- | --- | --- | --- | --- | --- |
| Total T cells | | 73.4 | 60.05-74.08 | 1695.5 | 1424-2664 |
|  | CD8+T cells | 34.9 | 19.68-34.06 | 807.1 | 518-1125 |
|  | CD8+ Naïve T cells | 38.7 | 41.58-77.90 | 312.4 | 297-730 |
|  | CD8+ TEMRA T cells | 21.7 | 1.70-24.62 | 175.1 | 11-218 |
|  | CD8+ CM | 21.4 | 12.08-30.54 | 172.7 | 85-268 |
|  | CD8+ EM | 18.2 | 1.58-13.18 | 146.9 | 10-129 |
|  | CD4+T cells | 34.9 | 26.17-40.76 | 807.1 | 686-1358 |
|  | CD4+ Naive T cells | 56.5 | 45.56-75.28 | 456.0 | 321-972 |
|  | CD4+ TEMRA | 0.6 | 0.00-1.06 | 5.0 | 0-13 |
|  | CD4+ CM | 33.2 | 22.06-46.46 | 268.0 | 211-478 |
|  | CD4+ EM | 9.7 | 2.08-8.78 | 78.2 | 23-84 |
|  | TCRαβ^+^DNT | 1.2 | 0.18-2.81 | 20.3 | 4-55 |
|  | γδ T cells | 8.0 | 6.92-19.84 | 136.3 | 124-410 |
| Total B cells | | 18.4 | 10.21-20.12 | 425.0 | 280-623 |
|  | Memory B | 4.5 | 7.76-19.90 | 19.1 | 31-94 |
|  | Naïve B | 90.2 | 48.36-75.84 | 383.4 | 147-431 |
|  | Transitional B | 5.2 | 2.58-12.30 | 21.9 | 10-66 |
|  | Plasmablasts B | 2.9 | 0.90-7.36 | 12.2 | 4-28 |
| NK cells | | 8.1 | 9.00-22.24 | 186.9 | 258-727 |
| CD4:CD8 | | 1 | 0.87-1.94 |  |  |

CM: central memory; EM: effector memory.

Table 2 The prediction of the identified variant in *ELF4*

| Gene name | Position | Transcript | Substitution | Polyphen2 | SIFT | MutationTaster | MutationAssessor | GERP++ |
| --- | --- | --- | --- | --- | --- | --- | --- | --- |
| ELF4 | chrX: 129205133 | NM_001421 | c.691T>C (p.W231R) | 0.906  (Probably Damaging) | 0.913  (Damaging) | 0.81  (Disease causing) | 0.933  (high impact) | 5.45 (Conserved) |
